# Increased Risk of Heart Rhythm Abnormalities in Adolescents and Young Adults who Vape: A Retrospective Cohort Study

**DOI:** 10.64898/2026.03.12.26348232

**Authors:** Athanasios Tsalatsanis, Joyce Johnson, Obada Abou-Assali, Shone Almeida, Racha Khalaf, Sami F. Noujaim

## Abstract

**Background:** Vaping among adolescents and young adults (AYA) could affect cardiovascular health. While pulmonary outcomes of vaping are well-documented, the link between vaping and abnormalities of heart rhythm remains unclear. We conducted a retrospective cohort study to test the hypothesis that the risk of heart rhythm abnormalities is increased in AYA who vape.

**Methods:** We used data from the TriNetX network to identify two cohorts of AYA (11 to 24 years old). The first cohort included individuals who vaped, and the second cohort was a comparison of individuals who did not report vaping. Individuals in the vaping cohort were matched 1:1 with those in the comparison cohort using propensity scores. The primary outcome was the association between vaping and diagnoses of heart rhythm abnormalities. The study analyzed data from 114,404 patients (57,202 in each cohort) with no significant differences in baseline characteristics.

**Results:** Patients who vaped had 82% higher odds of being diagnosed with heart rhythm abnormalities compared to those who did not vape (OR: 1.82, 95% CI: 1.74-1.91, p < 0.001). Furthermore, the hazard of developing heart rhythm abnormalities was approximately twice as high among vapers compared to those who did not (HR: 1.97, 95% CI: 1.88-2.06, p < 0.001).

**Conclusion:** This study shows a significant association between e-cigarette use and an increased risk of heart rhythm abnormalities in AYA. These findings highlight the potentially harmful cardiac electrophysiological outcomes of vaping in this population and underscore the importance of health interventions and surveillance.

## BACKGROUND

Vaping, or the use of electronic cigarettes (e-cigarettes), is a prevalent behavior specifically among adolescents and young adults (AYA) (1-6). For instance, it is estimated by the CDC that more than 2 million middle and high school students vape (2). Investigations into the health risks of vaping have centered on pulmonary outcomes and resulted in the identification of e-cigarette and vaping-associated lung injury (EVALI) as a distinct pathophysiological condition (7-14). However, the potential cardiovascular effects of e-cigarette use remain less understood, and concerns are growing about the undefined impact of vaping on cardiac arrhythmias, particularly in AYA.

E-cigarettes generate an inhalable aerosol, or e-vapor, by heating e-liquids that contain nicotine, propylene glycol, glycerin, and various flavoring agents. The harmful cardiovascular effects of nicotine, including in the pathogenesis of heart rhythm abnormalities, are well established in the context of traditional combustible cigarette smoking (15-21). Yet, the potential arrhythmogenic effects of inhaling e-cigarette vapor that contains aerosolized nicotine and the other chemicals present in e-liquids as well as their byproducts due to heating are not fully understood. Preclinical and clinical studies are pointing to possible arrhythmogenicity of vaping (13, 14, 22-25). Studies in rodents have suggested that vaping may lead to structural (26-29) and electrophysiological changes in the heart including decreased heart rate variability and increased inducibility of ventricular tachyarrhythmias (29-33). Clinical studies found that vaping increases the Tp-e/QT interval on ECG, which is linked to a higher risk of adverse cardiac events, including sudden death (34, 35). Clinical case studies further suggest possible association between vaping and heart rhythm abnormalities (36). A retrospective analysis of Mayo Clinic cases found a temporal link between vaping and sudden cardiac arrest or death, especially among younger individuals (37). Therefore, given the popularity of e-cigarettes among AYA, this population may be particularly vulnerable to the long-term cardiovascular consequences of vaping, and thus further investigations into the arrhythmogenic potential of vaping in this demographic are warranted.

Heart rhythm abnormalities, which can range from benign to life-threatening, may arise as a result of complex interactions between genetic susceptibility, environmental exposures, and lifestyle factors. In the context of vaping, the potential for e-cigarette use to influence the risk of abnormal heart rhythms among AYA, remains uncertain where vaping may act as an environmental exposure. The long-term cardiovascular implications due to such exposure are not fully understood given the relatively recent emergence of e-cigarettes.

In this retrospective cohort study, we aim to assess the prevalence of abnormal heart rhythms diagnoses among AYA who vape, in comparison to those who do not engage in vaping. Utilizing data from TriNetX, a federated research data network, we seek to determine whether vaping is associated with an increased risk of abnormal heart rhythms in this demographic. The findings from this study could provide insights into the potential cardiovascular risks of e-cigarette use, specifically in relation to arrhythmogenesis, and may contribute to the growing body of evidence on the adverse health consequences of vaping.

## METHODS

### Design and Setting

This is a retrospective cohort study using data from the TriNetX federated health research network. The TriNetX web-platform provides access to de-identified electronic health records of more than 150 million patients from 106 healthcare institutions across the United States. The records include diagnoses, procedures, medications, and laboratory results and are searchable via standardized terminologies. The TriNetX database captures longitudinal patient-level data, allowing for the follow-up of individuals over time across various inpatient and outpatient healthcare encounters. As such, it enables the assessment of temporal associations between exposures, diagnoses, treatments, and outcomes. This study is not considered human subjects research because the data are de-identified and the study is retrospective.

### Participants

We identified AYA (11 to 24 years old), who interacted with a healthcare system between January 1^st^, 2020 and January 1^st^, 2025. These subjects were classified into two cohorts: the vaping cohort (cases) and the non-vaping cohort (comparison) using ICD-10 codes. The cases included all patients with a diagnosis of a vaping related disorder or unspecified nicotine dependence during the study period. The comparison, non-vaping cohort did not have these diagnoses. We excluded patients with nicotine dependence attributed to cigarettes or tobacco products, other stimulant related disorders, history of congenital malformations of the heart, heart failure, cardiomyopathy, cardiac arrest, hypothyroidism, chronic kidney disease, kidney failure, attention-deficit hyperactivity disorders, and thyrotoxicosis. We also excluded patients prescribed with beta blockers, ace inhibitors, and calcium channel blockers.

The index event for the vaping cohort was the first diagnosis of vaping related disorder or unspecified nicotine dependence and for the comparison group, it was the first ambulatory visit in the study period. Patients in the vaping cohort were matched 1:1 with patients in the comparison cohort based on the demographic characteristics (i.e., age at index, gender, race, and ethnicity) and medication prescriptions (i.e., central nervous system medications, hormones/synthetics/modifiers, antihistamines, and respiratory tract medications) using scores. Baseline factors were assessed up to 6 months prior to the index event. Participants were followed for 5 years after the index event.

### Variables

Vaping was identified using the U07.0 and F17.20 ICD-10 codes. The primary outcome is the association between vaping and the development of heart rhythm abnormalities defined as the combination of the following diagnoses: other cardiac arrhythmias (ICD-10: I49), abnormalities of the heart beat (ICD-10: R00), or supraventricular tachycardia (ICD-10: I47.1). The complete list of the ICD-10 and CPT codes used to apply the exclusion criteria is included in the supplementary material. Other variables included demographics, and baseline medications.

### Bias and study size

All eligible vaping patients were included and matched 1:1 to controls using propensity scores.

### Statistical methods

The propensity scores were calculated based on demographics (i.e., age at index, gender, ethnicity, race) and medication use (central nervous system, hormones/synthetics,/modifiers, antihistamines, and respiratory tract medications) using logistic regression. Matching was performed using greedy nearest neighbor algorithms with a caliper width of 0.1 pooled standard deviations. At baseline, differences in continuous variables (i.e., age at index) in the matched cohorts were assessed using t-tests and differences in dichotomous outcomes (i.e., gender, race, etc.) were assessed with the Chi-square test. The primary outcome, the association between vaping and heart rhythm abnormalities, was evaluated both as a cross-sectional measure of diagnosis and as a time-to-event outcome. As a cross-sectional measure, it was assessed using Chi-square tests and summarized as odds ratios (OR) with 95% confidence intervals (CI). As a time-to-event outcome, it was assessed using Kaplan-Meier survival analysis. The difference in the distribution of the time to event outcome between the cohorts was assessed using the Log-Rank test. We report hazard ratios (HR) and 95% CI estimated from Cox proportional hazards models. Follow-up time was summarized using median and interquartile range (IQR). For ease of interpretation, we plot the cumulative incidence of heart rhythm abnormalities in both cohorts. Subgroup analyses assessed the association of vaping and heart rhythm abnormalities in patients 11 to 18 years old and in patients 19 to 24 years old. Further, to assess the robustness of the definition of heart rhythm abnormalities as a composite of other cardiac arrhythmias (ICD-10: I49), abnormalities of the heart beat (ICD-10: R00), or supraventricular tachycardia (ICD-10: I47.1), we conducted a sensitivity analysis restricting the definition of heart rhythm abnormalities to the I49 codes only. Statistical significance was set at p-value <0.05.

## RESULTS

### Participants

Out of 9,261,074 who met the inclusion criteria, 57,202 patients were vaping (cases) and 9,203,872 patients were not (comparison). After matching, 57,202 patients remained in each cohort. The demographic characteristics and medication prescriptions at baseline of the matched cohorts are presented in Table 1. There were no statistically significant differences in age, gender, race, ethnicity, or medication prescriptions between the two cohorts.

**Table 1:** Baseline characteristics of the matched cohorts.

|  | Vaping<br>(N=52702) | Comparison<br>(N=52702) | p-value |
| --- | --- | --- | --- |
| **Demographics** |  |  |  |
| Age at Index, Mean(SD) | 20.7 (2.57) | 20.7 (2.57) | 1 |
| Male | 30393 (53.13%) | 30393 (53.13%) | 1 |
| White | 31050 (54.28%) | 31050 (54.28%) | 1 |
| Black or African American | 12668 (22.15%) | 12668 (22.15%) | 1 |
| Hispanic or Latino | 7173 (12.54%) | 7172 (12.54%) | 0.993 |
| **Medications** |  |  |  |
| Central nervous system medications | 12088 (21.13%) | 12089 (21.13%) | 0.994 |
| Respiratory tract medications | 7243 (12.66%) | 7242 (12.66%) | 0.993 |
| Hormones/synthetics/modifiers | 6737 (11.78%) | 6737 (11.78%) | 1 |
| Antihistamines | 3915 (6.84%) | 3914 (6.84%) | 0.997 |

### Outcomes

The median follow-up was 210 days (IQR 0-810) in the cases cohort and 280 days (IQR:0-980) in the comparison cohort. There were 5,017 (8.77%) patients with heart rhythm abnormalities in the cases and 2,862 (5%) in the comparison.

### Main result

Vapers had 82% higher odds of being diagnosed with heart rhythm abnormalities compared to those who did not vape (OR: 1.82, 95% CI: 1.74-1.91, p < 0.001). Furthermore, the hazard of developing heart rhythm abnormalities was approximately twice as high among patients who vaped compared to those who did not (HR: 1.97, 95% CI: 1.88-2.06, p < 0.001).

### Subgroup analyses

#### Patients 11 to 18 years old

Each cohort included 10,695 matched patients. There were no statistically significant differences in baseline characteristics between the two cohorts (supplementary material). The median follow-up was 270 days (IQR: 0-820) in cases and 380 (IQR: 20-1100) in the comparison group. There were 1,027 (9.60%) patients with heart rhythm abnormalities in the cases group versus 531 (4.96%) in the comparison group. Vapers had 2 times higher odds of being diagnosed with heart rhythm abnormalities compared to non-vapers (OR: 2.03, 95% CI: 1.82-2.26, p < 0.001). The hazard of developing heart rhythm abnormalities was more than twice as high among vapers compared to those who did not (HR: 2.28, 95% CI: 2.05-2.54, p < 0.001).

#### Patients 19 to 24 years old

Each cohort included 47,086 matched patients. There were no statistically significant differences in baseline characteristics (supplementary material) between the two cohorts. The median follow-up was 200 days (IQR: 0-810) in cases and 280 (IQR: 0-920) in the comparison group. There were 4,047 (8.59%) patients with heart rhythm abnormalities in the cases cohort compared to 2,227 (4.73%) in the comparison cohort. Vapers had 89% higher odds of being diagnosed with heart rhythm abnormalities compared to those who did not vape (OR: 1.89, 95% CI: 1.79-2.00, p < 0.001). The hazard of developing heart rhythm abnormalities was twice as high among patients who vaped compared to those who did not (HR: 2.00, 95% CI: 1.90-2.11, p < 0.001).

### Sensitivity analyses

Patients who vaped had 59% higher odds of being diagnosed with heart rhythm abnormalities compared to non-vapers (OR: 1.59, 95% CI: 1.43-1.75, p < 0.001), and a 75% higher hazard of developing heart rhythm abnormalities over time (HR: 1.75, 95% CI: 1.58-1.94, p < 0.001). In patients 11-18 years old, vaping was associated with 78% higher odds of heart rhythm abnormalities diagnosis (OR: 1.78, 95% CI: 1.43-2.21, p < 0.001) and a two-fold higher hazard of incident heart rhythm abnormalities (HR: 2.08, 95% CI: 1.68-2.59, p < 0.001). In patients 19-24 years old, vaping was associated with 73% higher odds of heart rhythm abnormalities diagnosis (OR: 1.73, 95% CI: 1.54-1.94, p < 0.001) and an 85% higher hazard of incident heart rhythm abnormalities (HR: 1.85, 95% CI: 1.65-2.08, p < 0.001).

## DISCUSSION

Previous studies have mainly focused on the pulmonary consequences of e-cigarette use, such as EVALI (7-14). In this TriNetX retrospective cohort study of AYA aged 11 to 24, there was a statistically significant association between vaping and the incidence of heart rhythm abnormalities. Vapers were twice as likely to develop heart rhythm abnormalities compared to their non-vaping cohorts. These findings suggest that vaping may negatively impact cardiac electrophysiology, even in relatively young and healthy individuals.

The potential proarrhythmogenic cardiovascular impact, such as endothelial dysfunction, increased sympathetic activation, and alterations in heart rate variability has been investigated in prior studies (24, 25, 29, 33-35, 38-44). Our findings here align with these prior studies and indicate that vaping is associated with clinically diagnosed heart rhythm disturbances in a real-world cohort (36, 37).

Mechanistically, the association between heart rhythm abnormalities and vaping may be due in part to nicotine, an important constituent of most e-liquids, which has well-established pro-arrhythmic properties, including increased catecholamine release, prolonged QT interval, and altered myocardial excitability (13, 15-21, 31). Additionally, other constituents of e-cigarette vapor such as aldehyde byproducts of heating, particulate matter, and flavoring agents have been implicated in oxidative stress and inflammatory responses that could also contribute to structural and electrical remodeling of the myocardium (30, 44-58).

The strengths of this study include using a large database that can query the de-identified electronic health records of more than 150 million patients from 106 healthcare institutions across the United States across inpatient and outpatient settings, and propensity score matching to control for baseline differences. On the other hand several limitations are present: the observational design precludes the ability to establish causality, the reliance on ICD codes which may introduce misclassification bias, and our current analysis based on ICD data does not allow to differentiate the types of arrhythmias present, the unmeasured confounders such as illicit substance use, duration and frequency of vaping, the unknown specifics of the vaping product used (disposable or refillable systems, nicotine salt, or nicotine free base, and the concentration of nicotine), and the other environmental or genetic factors that may predispose to arrhythmias which could not be accounted for. Additionally, it should be noted that the median follow-up period was relatively short, and long-term cardiovascular risks need to be evaluated based on appropriately designed longitudinal studies. Finally, we wish to point out that while it is possible that vaping is more often coded in those who have potential harm related to vaping, the diagnoses of heart rhythm abnormalities are not likely to be reported more often due to a vaping diagnosis; consequently, the finding of a correlation with vaping and heart rhythm abnormalities remains valid. This correlation requires additional hypothesis testing to determine the clinical significance of our findings.

## CONCLUSONS

Given the high prevalence of vaping among adolescents and young adults, the observed association with arrhythmic events in this population highlights an increased need for targeted public health interventions, tailored prevention efforts and continued surveillance. Our findings underscore the need for improved clinical awareness and further basic, translational, clinical and longitudinal studies into pathophysiological mechanisms and short and long-term cardiovascular effects of e-cigarette use in younger populations.

## Data Availability

The data that support the findings of this study are available from TriNetX Research Network but restrictions apply to the availability of these data, which were used under license for the current study, and so are not publicly available. Data are however available from the authors upon reasonable request and with permission of TriNetX.

## LIST OF ABBREVIATIONS

AYA: adolescents and young adults
CI: confidence intervals
e-cigarettes: electronic cigarettes
EVALI: e-cigarette and vaping-associated lung injury
HR: hazard ratios
IQR: interquartile range
OR: odds ratio

## DECLARATIONS

### Ethics approval and consent to participate

Not applicable. This study is not considered human subjects research because the data are de-identified and the study is retrospective.

### Consent for publication

Not applicable

## Authors’ contributions

Dr. Athanasios Tsalatsanis, Dr. Racha Khalaf, and Dr. Sami Noujaim conceptualized and designed the study, drafted the initial manuscript, and critically reviewed and revised the manuscript. Dr. Joyce Johnson, Dr. Shone Almeida, and Obada Abou-Assali drafted the initial manuscript, and critically reviewed and revised the manuscript. All authors approved the final manuscript as submitted and agree to be accountable for all aspects of the work.

## Acknowledgements

Not applicable

## Authors’ information

Not applicable

## LEGENDS

Supplementary material Table A1: ICD-10 and CPT Codes used to define the exclusion criteria for the population of this study.

Supplementary material Table A2: Baseline characteristics of the matched cohorts.

Supplementary material Table A3: Baseline characteristics of the matched cohorts.

