## Appendix for "Increased Risk of Heart Rhythm Abnormalities in Adolescents and Young Adults who Vape: A Retrospective Cohort Study"

Supplementary Material

**METHODS**

**Variables**

**Table A1.** ICD-10 and CPT Codes used to define the exclusion criteria for the population of this study

| Cases | |
| --- | --- |
| Diagnoses, ICD-10 Code | Description |
| Q24 | Other congenital malformations of heart |
| F90 | Attention-deficit hyperactivity disorders |
| F15 | Other stimulant related disorders |
| E05 | Thyrotoxicosis [hyperthyroidism] |
| E03 | Other hypothyroidism |
| I42 | Cardiomyopathy |
| I50 | Heart failure |
| N18 | Chronic kidney disease |
| N19 | Unspecified kidney failure |
| I46 | Cardiac arrest |
| Z72.0 | Tobacco use |
| F17.21 | Nicotine dependence, cigarettes |
| F17.29 | Nicotine dependence, other tobacco product |
| Procedures, CPT code |  |
| 1012740 | Dialysis Services and Procedures |
| Medications, TriNetX Codes |  |
| CV100 | Beta blockers |
| CV800 | Ace inhibitors |
| CV200 | Calcium channel blockers |
| Comparison | |
| Diagnoses, ICD-10 Code | Description |
| Q24 | Other congenital malformations of heart |
| F90 | Attention-deficit hyperactivity disorders |
| F15 | Other stimulant related disorders |
| E05 | Thyrotoxicosis [hyperthyroidism] |
| E03 | Other hypothyroidism |
| I42 | Cardiomyopathy |
| I50 | Heart failure |
| N18 | Chronic kidney disease |
| N19 | Unspecified kidney failure |
| I46 | Cardiac arrest |
| Z72.0 | Tobacco use |
| F17 | Nicotine dependence |
| Z87.891 | Personal history of nicotine dependence |
| U07.0 | Vaping-related disorder |
| Procedures, CPT code |  |
| 1012740 | Dialysis Services and Procedures |
| Medications, TriNetX Codes |  |
| CV100 | Beta blockers |
| CV800 | Ace inhibitors |
| CV200 | Calcium channel blockers |

**RESULTS**

**Subgroup analyses**

**Participants 11 to 18 years old**:

**Table A2.** Baseline characteristics of the matched cohorts.

|  | **Vaping (N=10695)** | **Comparison (N=10695)** | **p-value** |
| --- | --- | --- | --- |
| **Demographics** |  |  |  |
| Age at Index, Mean(SD) | 16.62 (1.60) | 16.62 (1.60) | 0.993 |
| Male | 5106 (47.74%) | 5104 (47.72%) | 0.978 |
| White | 6137 (57.38%) | 6137 (57.38%) | 1 |
| Black or African American | 2064 (19.30%) | 2062 (19.28%) | 0.972 |
| Hispanic or Latino | 1608 (15.04%) | 1610 (15.05%) | 0.969 |
| **Medications** |  |  |  |
| Central nervous system medications | 2794 (26.12%) | 2794 (26.12%) | 1 |
| Respiratory tract medications | 1728 (16.16%) | 1728 (16.16%) | 1 |
| Hormones/synthetics/modifiers | 1625 (15.19%) | 1627 (15.21%) | 0.969 |
| Antihistamines | 1058 (9.89%) | 1058 (9.89%) | 1 |

**Participants 19 to 24 years old**:

**Table A3.** Baseline characteristics of the matched cohorts.

|  | **Vaping (N=47086)** | **Comparison (N=47086)** | **p-value** |
| --- | --- | --- | --- |
| **Demographics** |  |  |  |
| Age at Index, Mean(SD) | 21.68 (1.67) | 21.68 (1.67) | 0.998 |
| Male | 25571 (54.31%) | 25571 (54.31%) | 1 |
| White | 25218 (53.56%) | 25219 (53.56%) | 0.994 |
| Black or African American | 10755 (22.84%) | 10754 (22.84%) | 0.993 |
| Hispanic or Latino | 5636 (11.97%) | 5636 (11.97%) | 1 |
| **Medications** |  |  |  |
| Central nervous system medications | 9503 (20.18%) | 9504 (20.18%) | 0.993 |
| Respiratory tract medications | 5661 (12.02%) | 5661 (12.02%) | 1 |
| Hormones/synthetics/modifiers | 5235 (11.12%) | 5235 (11.12%) | 1 |
| Antihistamines | 2936 (6.24%) | 2936 (6.24%) | 1 |
